# Epidemiological and clinical characteristics of discharged patients infected with SARS-CoV-2 on the Qinghai plateau

**DOI:** 10.1101/2020.04.23.20077644

**Authors:** Aiqi Xi, Ma Zhuo, Jingtao Dai, Yuehe Ding, Xiuzhen Ma, Xiaoli Ma, Xiaoyi Wang, Lianmeng Shi, Huanying Bai, Hongying Zheng, Eric Nuermberger, Jian Xu

## Abstract

Since the outbreak of coronavirus disease 2019 (COVID-19), caused by the severe acute respiratory syndrome coronavirus 2 (SARS-CoV-2) was first reported in Wuhan, a series of confirmed cases of COVID-19 were found on the Qinghai-Tibet plateau. We aimed to describe the epidemiological, clinical characteristics, and outcomes of all confirmed cases in Qinghai, a province at high altitude. With efficient measures to stop the spread of coronavirus, no new cases were found in Qinghai Province for 60 consecutive days between Feb 6 and April 6, 2020. Of all 18 patients with confirmed SARS-CoV-2 infection, 15 patients comprising 4 transmission clusters were identified. Three patients were infected by direct contact without travel history to Wuhan. Seven patients were asymptomatic on admission. Of 18 patients, 10 patients showed bilateral pneumonia and 2 patients showed no abnormalities. Three patients with comorbidities such as hypertension, liver diseases or diabetes developed severe illness. High C-reactive protein levels and elevations of both ALT and AST were observed in 3 severely ill patients on admission. All 18 patients were eventually discharged, including the 3 severe patients who recovered after treatment with non-invasive mechanical ventilation, convalescent plasma and other therapies. Our findings confirmed human-to-human transmission of SARS-CoV-2 in clusters. The strategies of early diagnosis, early isolation, and early treatment are important to prevent the spread of COVID-19 and improve the cure rate. Patients with comorbidities are more likely to develop severe illness and could benefit from convalescent plasma transfusion.

## Introduction

Coronavirus disease 2019 (COVID-19), caused by infection with the novel severe acute respiratory syndrome coronavirus 2 (SARS-CoV-2), emerged in Wuhan, Hubei, China in December 2019^1-4^ and rapidly spread worldwide.^5^ The outbreak spread to 209 countries and the global number of reported cases surpassed 1,210,000 as of April 6, 2020.^6^ With evidence that SARS-CoV-2 is spread by human-to-human transmission,^7-9^ the increasing number of cases and widening geographical spread of the disease raise a global health concern.^10^

So far, several studies have described the epidemiological and clinical features of COVID-19, but the data mainly came from Wuhan.^4,11^ Qinghai province, located on the Qinghai-Tibet plateau with an average altitude of more than 3000 meters above sea level and a population of 6.03 million, reported a total of 18 confirmed cases by April 6. During the outbreak, Qinghai rapidly instituted a number of strict control measures to lower transmission, including the enforcement of quarantine measures, early detection, reducing passenger flow, and strong social messaging. By April 6, 2020, no new confirmed cases had been found in Qinghai Province for 60 consecutive days since Feb 6, 2020. More importantly, all 18 patients including 3 severely ill cases had been discharged after treatment by Feb 21, 2020 (Figure 1).

**Figure 1:**
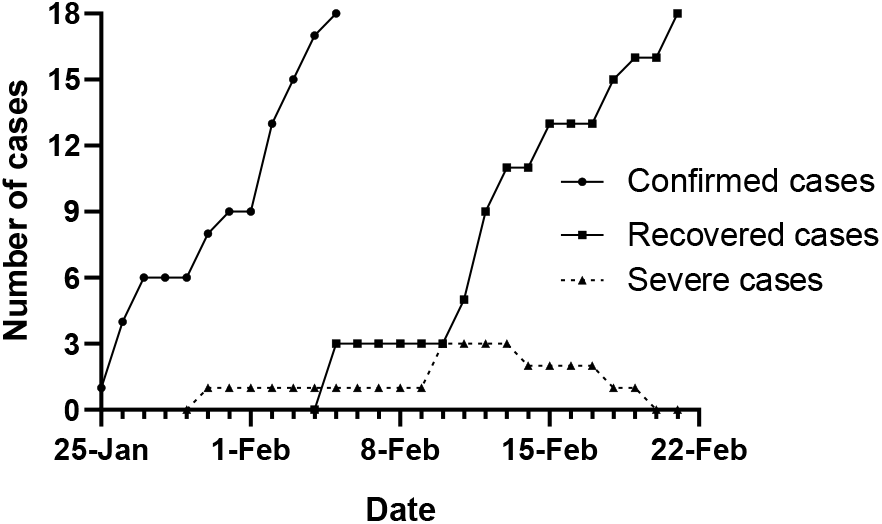
Epidemic curves of all patients with confirmed SARS-CoV-2 infection in Qinghai, China.

In this study, we report the epidemiological and clinical characteristics, and outcomes of all 18 confirmed COVID-19 patients in Qinghai including family clusters who returned to Qinghai from Wuhan, and family members who did not travel to Wuhan.

## Methods

### Study design and participants

For this retrospective study, we enrolled all 18 patients infected with SARS-CoV-2 from the hospitals designated for treatment by the Health Commission of Qinghai Province from Jan 21 to April 6, 2020. 15 patients were from the Fourth People’s Hospital of Qinghai Province and 3 patients were from the Third People’s Hospital of Xining. All confirmed patients enrolled in this study were diagnosed with COVID-19 according to World Health Organization interim guidance^12^. A confirmed case of COVID-19 was defined as a positive result on real-time reverse-transcriptase polymerase chain reaction (RT-PCR) assay from nasal and pharyngeal swab specimens. Incubation period was defined from contact with a symptomatic case to illness onset. This study was approved by the ethics commissions of the two hospitals. The requirement for informed patient consent was waived by the ethics committees for this retrospective study.

### Data collection

The epidemiological, demographic, clinical, laboratory and radiological characteristics and treatment and outcomes data were obtained from patients’ medical records. Information recorded included demographic data, exposure history, comorbidities, symptoms, laboratory findings and computed tomographic (CT) scans. Laboratory confirmation of SARS-CoV-2 by real-time PCR was done in Qinghai Center for Diseases Prevention and Control. The severity of COVID-19 was defined based on COVID-19 Guidelines (5th version) made by the National Health Commission of the People’s Republic of China (NHCC). Qinghai is located on a plateau, with an average altitude of 2261 meters above sea level in Xining area. The atmospheric pressure and air oxygen content are low. Therefore, oxygenation index should be calculated as follows: PaO2/[FIO2×(barometric pressure/760)] according to the Berlin Definition of Acute Respiratory Distress Syndrome^13^.

Mild cases: clinical symptoms were mild without pneumonia manifestation through image results. Moderate cases: having fever and other respiratory symptoms with pneumonia manifestation through image results. Severe cases: meeting any one of the following: respiratory distress, RR> 30/min; SpO2 ≤90% at rest in Xining adjusted according to altitude; PaO2/FiO2≤300mmHg needed to be corrected according to altitude as mentioned above.

All treatment measures were collected during the hospitalization, such as antiviral therapy, antibacterial therapy, corticosteroid therapy, traditional Chinese medicine therapy, immune support therapy, convalescent plasma therapy, and respiratory support. Discharge criteria were based on COVID-19 Guidelines (5th version) by NHCC as follows: body temperature normal for more than 3 days, respiratory symptoms and pulmonary imaging improved significantly, and respiratory tract specimen nucleic acid amplification test negative on two consecutive occasions at least 24 hours apart.

### Statistical analysis

Categorical variables were described as frequency rates and percentages, and continuous variables were described using median and interquartile range (IQR) values. Statistical analyses were done using the Graphpad Prism software, version 8.02, unless otherwise indicated.

## Results

### Epidemiological analysis

All 18 patients identified as confirmed SARS-CoV-2 cases were included in this study from January 25 to February 5, 2020 (Figure 1). Among them, 3 (17%), 13 (72%) and 2 (11%) patients were categorized into severe, moderate and mild groups, respectively, during hospitalization. Of 3 severe patients, 2 were initially classified as moderate and then changed to severe as disease progressed. In total, 15 patients returned from Wuhan, Hubei Province of China. Of the 3 remaining cases, 2 patients had contact with a confirmed case and 1 patient had contact with a family member who returned from Wuhan with a negative result on nucleic acid amplification test. In total, 4 clusters of SARS-CoV-2 infection were identified, involving 15 cases. The median age was 32 years (range, 7-47 years), and 12 (67%) were men. Five (28%) patients, including 3 severe patients, had chronic diseases, including hypertension, hyperlipidemia, diabetes, liver injury and polymyositis (Table 1 and 2). On admission, the most common symptoms were cough (9 [50%]), sputum production (6 [33%]), chest tightness (6 [33%]), fever (3 [17%]) and fatigue (3 [17%]). All 3 severe patients showed cough, sputum production and chest tightness but only 2 showed fever (Table 2). Less common symptoms were sore throat, and diarrhoea (Table 1). Three second-generation cases had credible information on contacts with incubation periods of 5, 14 and 15 days. The median time from departure from Wuhan to admission was 8 days (IQR, 2-12).

**Table 1:**
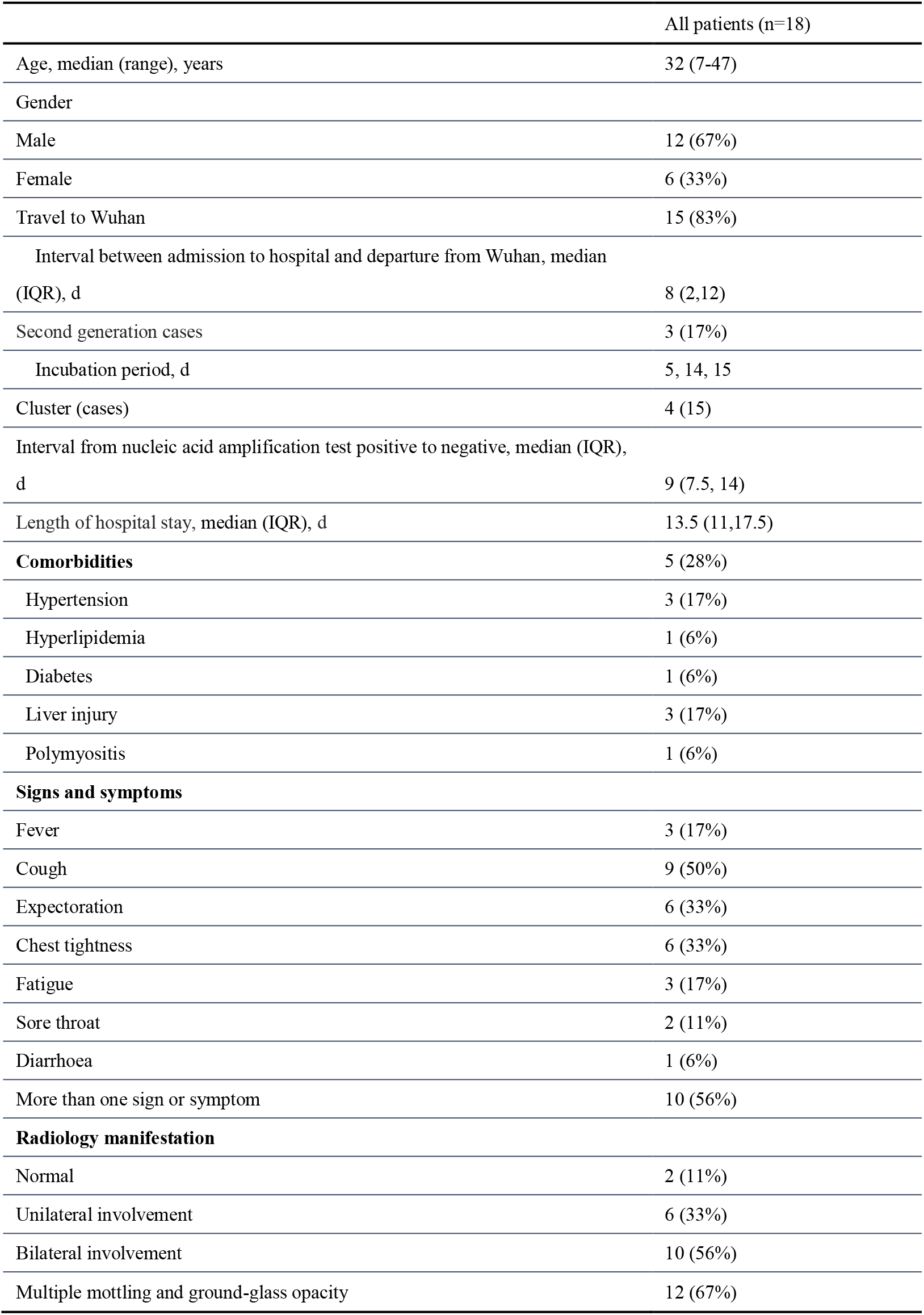
Demographics, comorbidities, clinical symptoms and radiology findings on admission.

|  | All patients (n=18) |
| --- | --- |
| Age, median (range), years | 32 (7-47) |
| Gender |  |
| Male | 12 (67%) |
| Female | 6 (33%) |
| Travel to Wuhan | 15 (83%) |
| Interval between admission to hospital and departure from Wuhan, median (IQR), d | 8 (2,12) |
| Second generation cases | 3 (17%) |
| Incubation period, d | 5, 14, 15 |
| Cluster (cases) | 4 (15) |
| Interval from nucleic acid amplification test positive to negative, median (IQR), d | 9 (7.5, 14) |
| Length of hospital stay, median (IQR), d | 13.5 (11,17.5) |
| **Comorbidities** | 5 (28%) |
| Hypertension | 3 (17%) |
| Hyperlipidemia | 1 (6%) |
| Diabetes | 1 (6%) |
| Liver injury | 3 (17%) |
| Polymyositis | 1 (6%) |
| **Signs and symptoms** |  |
| Fever | 3 (17%) |
| Cough | 9 (50%) |
| Expectoration | 6 (33%) |
| Chest tightness | 6 (33%) |
| Fatigue | 3 (17%) |
| Sore throat | 2 (11%) |
| Diarrhoea | 1 (6%) |
| More than one sign or symptom | 10 (56%) |
| **Radiology manifestation** |  |
| Normal | 2 (11%) |
| Unilateral involvement | 6 (33%) |
| Bilateral involvement | 10 (56%) |
| Multiple mottling and ground-glass opacity | 12 (67%) |

**Table 2:** Clinical characteristics of 3 severe patients with COVID-19.

|  | On admission |  |  |  |  |  | Interval<br>from onset<br>to severe<br>illness | Severe | Discharged |
| --- | --- | --- | --- | --- | --- | --- | --- | --- | --- |
|  | Comorbidities | symptoms | Radiology | ALT,<br>U/L | AST,<br>U/L | C-reactive<br>protein,<br>mg/L |  | Corrected<br>PaO2/FiO2 | length of<br>hospital stay |
| Case 1 | Hypertension,<br>hyperlipidemia,<br>diabetes, liver<br>injury | Cough, sputum<br>production,<br>chest tightness,<br>fatigue | Bilateral<br>pneumonia | 151 | 76 | >20 | 13 | 217 | 16 |
| Case 2 | Hypertension,<br>liver injury | Fever, cough,<br>sputum<br>production,<br>chest tightness,<br>fatigue | Bilateral<br>pneumonia | 78 | 64 | 53.92 | 8 | 174 | 18 |
| Case 3 | Hepatitis B | Fever, cough,<br>sputum<br>production,<br>chest tightness | Bilateral<br>pneumonia | 82 | 101 | 17.92 | 8 | 163 | 18 |

ALT= Alanine aminotransferase. AST= Aspartate aminotransferase. PaO2=partial pressure of oxygen. FiO2=fraction of inspired oxygen.

### Imaging features

Chest X-ray or CT examination was performed on all patients on admission. Of 18 patients, 10 (56%) patients showed bilateral pneumonia while 6 (33%) patients showed unilateral pneumonia, and 2 (11%) patients showed no abnormalities (Table 1). The most common abnormalities were ground-glass opacities (12 [67%]) and patchy shadows (Figure 2).

**Figure 2:**
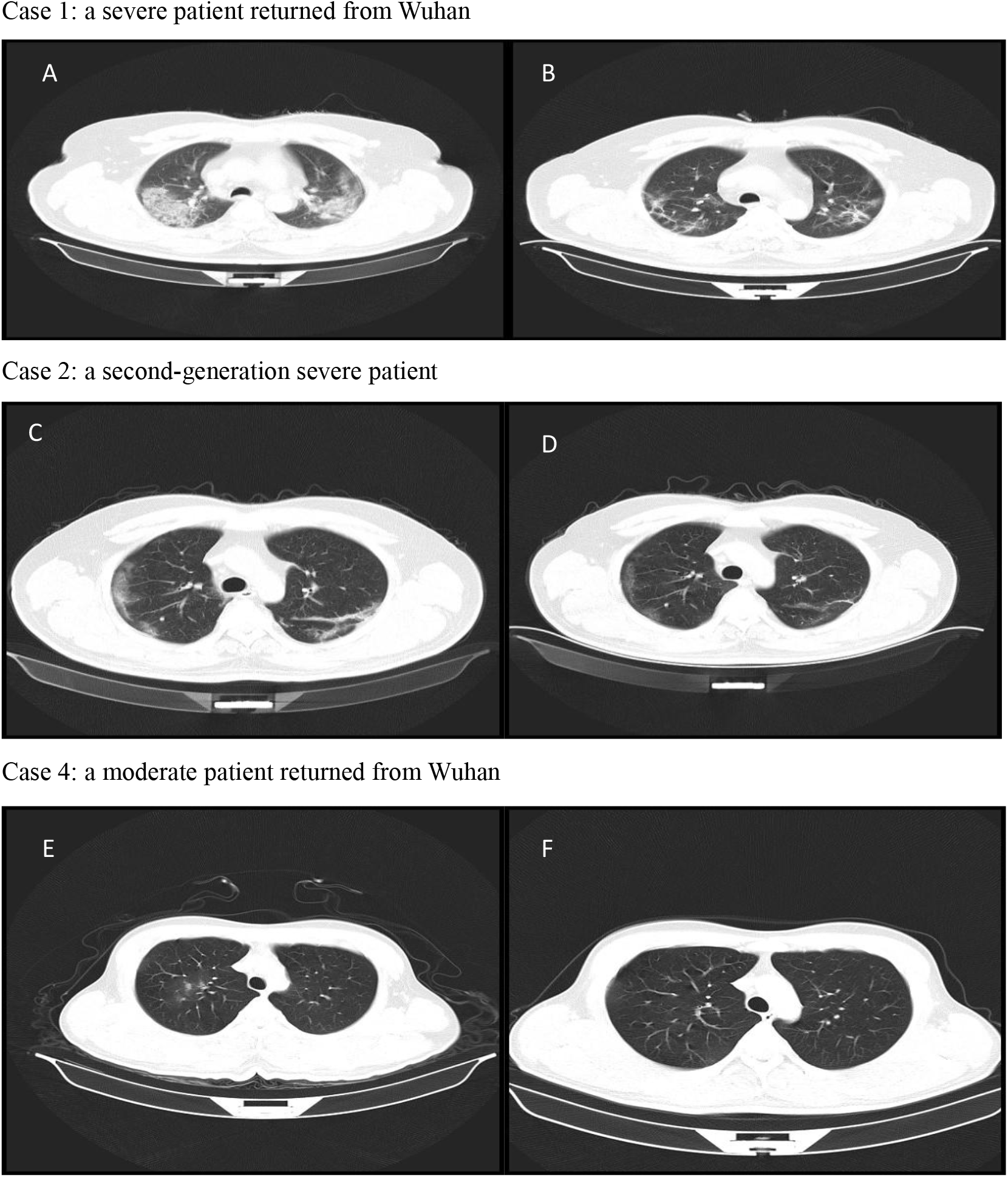
Chest computed tomographic (CT) images of patients infected with SARS-CoV-2. Case 1: A. CT scan showing mass shadows of consolidation and bilateral ground glass opacities on day 13 after symptom onset; B. Image showing the resolution of bilateral ground glass opacities and a decrease of consolidation after treatment. Case 2: C. CT scan showing bilateral ground glass opacities on day 6 after symptom onset; D. Image showing the resolution of bilateral ground glass opacities and low density shadow after treatment. Case 4: E. CT scan showing ground glass opacities on day 8 after symptom onset; F. Image showing the resolution of the lesions after treatment.

### Laboratory findings

On admission, 3 (17%), 3 (17%), 4 (22%) and 5 (28%) patients had leucopenia, lymphopenia, neutropenia and eosinophilia, respectively (Table 3). Hemoglobin was above the normal range in 6 (33%) patients, which may be attributed to high altitude. Both alanine aminotransferase (ALT) and aspartate aminotransferase (AST) concentrations were elevated in 4 patients including 3 severe patients (Table 2). Elevated levels of lactate dehydrogenase (LDH) and creatine kinase were found in 6 (40%) and 2 (13%) cases, respectively. One patient with polymyositis showed abnormal creatine kinase (3510 U/L), LDH (527 U/L), ALT (82 U/L) and AST (101 U/L). Elevations of glucose and lactic acid were found in about 44%. Similarly, 7/16 and 5/16 patients demonstrated elevated C-reactive protein levels and erythrocyte sedimentation rates, respectively. Notably, the levels of C-reactive protein in 3 severe patients were higher than those of mild and moderate patients (Table 2 and 3). Levels of D-dimer and procalcitonin were in the normal range in 12 detected patients.

**Table 3:** Laboratory examination of patients infected with SARS-CoV-2 on admission.

|  | Normal range | Median (IQR) | Proportion of cases with higher than normal value | Proportion of cases with lower than normal value |
| --- | --- | --- | --- | --- |
| **Blood routine and lymphocyte classification** |  |  |  |  |
| White blood cells, $\times 10^9/L$ | 4.00-10.00 | 4.8 (4.2,6.6) | 0/18 | 3/18 |
| Lymphocytes, $\times 10^9/L$ | 0.8-4 | 1.4 (1.1,1.8) | 0/18 | 3/18 |
| Neutrophils, $\times 10^9/L$ | 2-7.7 | 2.8 (2.1,3.5) | 0/18 | 4/18 |
| Eosinophils, $\times 10^9/L$ | 0.02-0.5 | 0.04 (0.01-0.08) | 0/18 | 5/18 |
| Hemoglobin, g/L | 110-160 | 156 (141,166) | 6/18 | 0 |
| Platelets, $\times 10^9/L$ | 100-300 | 171 (125,205) | 0/18 | 2/18 |
| **Blood biochemistry** |  |  |  |  |
| Alanine aminotransferase, U/L | 0-50 | 45 (22,65) | 6/15 | 0 |
| Aspartate aminotransferase, U/L | 0-50 | 28 (22,47) | 4/15 | 0 |
| Total bilirubin, $\mu\text{mol/L}$ | 0-22 | 10.5 (7.5,13.3) | 0/15 | 0 |
| Albumin, g/L | 35-55 | 43 (39,45) | 0/15 | 0 |
| Lactate dehydrogenase, U/L | 110-245 | 215 (176,276) | 6/15 | 0 |
| Creatine kinase, U/L | 25-200 | 65 (42,97) | 2/15 | 0 |
| Blood urea nitrogen, mmol/L | 1.7-8.6 | 5.0 (3.5,5.9) | 0/15 | 0 |
| Glucose, mmol/L | 3.8-6.2 | 6.1 (5.6,7.1) | 8/18 | 0 |
| Lactic acid, mmol/L | 0.5-1.6 | 1.6 (1.2,1.9) | 7/16 | 0 |
| Potassium, mmol/L | 3.5-5.5 | 3.8 (3.4,4.1) | 2/18 | 6/18 |
| Sodium, mmol/L | 135-145 | 138 (136,139) | 0/18 | 3/18 |
| **Coagulation function** |  |  |  |  |
| Prothrombin time,s | 10--15 | 12.9 (12.4,13.2) | 0/15 | 0 |
| Activated partial thromboplastin time, s | 22--40 | 33 (29,35) | 1/15 | 0 |
| Fibrinogen degradation products, $\mu\text{g/mL}$ | 0-5 | 1.2 (0.9,1.7) | 0/13 | 0 |
| D-dimer,$\mu\text{g/mL}$ | 0-1 | 0.24 (0.18,0.32) | 0/12 | 0 |
| **Infection-related parameters** |  |  |  |  |
| Procalcitonin, ng/mL | 0-0.15 | Normal | 0/12 | 0 |
| C-reactive protein, mg/L | 0-3 | 2.2 (0.3-12.8) | 7/16 | 0 |
| Erythrocyte sedimentation rate, mm/h | 0-20 | 13.5 (6.8,29.3) | 5/16 | 0 |

### Treatment

All 18 patients received antiviral treatment, including lopinavir/ritonavir (18 [100%]), interferon-α2b (18 [100%]), oseltamivir (5 [28%]), and ribavirin (6 [33%]) (Table 4). 17 patients initiated lopinavir/ritonavir and interferon-α2b treatment within one day after admission. The median duration of lopinavir/ritonavir and interferon-α2b treatment was 8 days (IQR, 5.5-10). 11 (61%) patients were given antibacterial treatment (moxifloxacin) and all patients were treated with various traditional Chinese medicines according to different symptoms and signs of each person. One patient with moderate COVID-19 showed cardiotoxicity with high level of creatine kinase (2756 U/L) after treatment with moxifloxacin and lopinavir/ ritonavir.

**Table 4:** Complications and treatments of all 18 patients infected with SARS-CoV-2.

|  | No. (%) |
| --- | --- |
| **Complications** |  |
| ARDS* | 3 (17%) |
| Liver injury* | 3 (17%) |
| **Treatment** |  |
| **Oxygen therapy** |  |
| Nasal cannula | 10 (56%) |
| Non-invasive ventilation or high-flow nasal cannula# | 3 (17%) |
| Mechanical ventilation (non-invasive)# | 3 (17%) |
| **Antiviral treatment** |  |
| Lopinavir/ritonavir, interferon-α2b | 18 (100%) |
| Oseltamivir | 5 (28%) |
| Ribavirin | 6 (33%) |
| Antibacterial treatment, moxifloxacin | 11 (61%) |
| Methylprednisolone# | 3 (17%) |
| Immunoregulation therapy, thymalfasin, immunoglobulin# | 3 (17%) |
| Convalescent plasma | 4 (22%) |
| Traditional Chinese medicine | 18 (100%) |

\*Complications occurred in 3 severe patients. #These therapies were only used for 3 severe patients.

10 (56%) patients received oxygen therapy by nasal cannula. The corrected PaO2/FiO2 values of 3 severe patients were 217, 174 and 163 mm Hg, respectively, which were below 300 mm Hg. During hospitalization, all 3 severe patients had complications including acute respiratory distress syndrome (ARDS), acute respiratory injury and liver injury. These 3 severe cases were given non-invasive mechanical ventilation for respiratory support. Besides antiviral therapy, antibacterial therapy and traditional Chinese medicine, all 3 severe patients received methylprednisolone (40 mg, bid, 3 days), immunoglobulin (20-25g, qd, 3 days), thymalfasin (0.4 mg, qod, 3 times), and convalescent plasma (50 ml, qod, twice) (Table 4).

### Outcomes

With treatment, the condition of all 18 patients improved significantly and CT images showed obvious regression of ground glass opacities (Figure 2). All patients including 3 severe cases were discharged. The median time for conversion of nucleic acid amplification test from positive to negative was 9 days (IQR, 7.5-14) and the median length of hospital stay was 13.5 days (IQR, 11-17.5).

## Discussion

The first laboratory-confirmed case of COVID-19 in Qinghai was reported on January 25, 2020 and the province leadership initiated first-level response on the same day. A total of 15 cases were imported from Wuhan before the city was sealed off on January 23. Overall, 4 transmission clusters involving 15 cases (2-5 cases/cluster) were identified. Three patients without history of travel to Wuhan were infected by direct contact with cases returned from Wuhan. These data clearly confirmed human-to-human transmission of COVID-19^7,8,14^ and the effectiveness of isolation to control the epidemic. Chinese and Qinghai governments issued very strict and efficient measures to stop the spread of SARS-CoV-2 such as joint prevention and control, mobility reduction, cancellation of gathering activities, enforcement of quarantine measures, and social messaging about personal protection. With the substantial public health prevention measures and huge efforts from medical professionals to treat patients, 18 patients were confirmed infected with SARS-CoV-2 from Jan 25 to Feb 5, 2020 and all of them were discharged between Feb 5 and Feb 21, 2020. A total of 437 close contacts of the confirmed cases were released after medical observation. By April 16, no new cases were found in Qinghai Province for 60 consecutive days since Feb 6, 2020. Notably, our medical workers had zero infection during this outbreak.

Due to the early detection and early diagnosis strategies, most patients were confirmed at early stage. Five symptomatic patients infected with SARS-CoV-2 were admitted within 3 days of returning from Wuhan. The 7 (39%) asymptomatic patients in our Qinghai cohort were diagnosed after nucleic acid amplification test and 2 of them had normal chest CT examinations. It is worth noting that one second-generation patient had contact with, and presumably contracted COVID-19 from, his son who returned from Wuhan with no symptoms. Therefore, to identify and contain the asymptomatic cases are the important measures to prevent transmission on the COVID-19.

We observed a greater number of men than women among the 18 cases of SARS-CoV-2 infection, consistent with a previous study^15^. Additionally, 3 children were infected with SARS-CoV-2 and showed mild or moderate symptoms. All 3 severe cases had comorbidities such as hypertension, liver disease or diabetes. SARS-CoV-2 infects host cells through angiotensin-converting enzyme 2 (ACE2) receptors.^2^ ACE2 is highly expressed in the heart and lungs, which is involved in heart function and the development of hypertension and diabetes.^16^ Liver injury in patients with SARS-CoV-2 infections might be also directly caused by the viral infection of liver cells.^17^ Elevation of both ALT and AST was observed in 4 patients including 3 severe cases and 1 case with polymyositis on admission. Therefore, liver damage is more prevalent in severe cases than in mild and moderate cases of COVID-19 consistent with previous reports.^4,14^ As the elevated amount of C-reactive protein may be associated with the inflammatory response and cytokine storms caused by the virus in the blood vessels ^18^, a previous study showed that the C-reactive protein level was positively correlated with the severity of the pneumonia.^19^ Similarly, we found that the amount of C-reactive protein was higher in 3 severe patients than the other 15 mild and moderate patients.

Qinghai is located on an elevated plateau with lower ambient oxygen levels. Compared to those living at lower altitudes, patients at high altitude are less tolerant to hypoxia and lung diseases are more likely to cause respiratory failure.^20^ Therefore, oxygen supply is important for patients with COVID-19, especially severe patients. Our 3 severe patients had acute respiratory distress syndrome (ARDS) with low PaO2/FiO2 prompting non-invasive mechanical ventilation. In addition to antiviral and antibiotic therapy and traditional Chinese medicine, all 3 severe cases received immunoglobulin and methylprednisolone for a short duration.

Convalescent plasma has been used to improve the survival rate of patients with severe acute respiratory syndrome (SARS) coronavirus infection.^21^ In 2015, the use of convalescent plasma for the treatment of Middle East Respiratory Syndrome (MERS) coronavirus was established as a protocol.^22^ Thus, our 3 severe patients were given convalescent plasma (50ml, qod, twice) collected from 2 patients who had recovered from COVID-19. We detected SARS-CoV-2 antibodies (IgG and IgM) from the convalescent plasma of one donated patient using chemiluminescent immunoassay. The level of IgG was very high (>30 AU/mL) and IgG (1:80) was 3.464 AU/mL. As expected, the level of IgM was very low (0.093 AU/mL). The CT images, blood gas analysis and symptoms improved after convalescent plasma transfusion. No adverse events were observed. One possible explanation for the efficacy of convalescent plasma is that the antibodies from convalescent plasma might suppress viraemia.^23^

Our study has several limitations. First, with the limited number of cases in Qinghai province, the results should be interpreted with caution. Second, we did not investigate the correlation between the viral load and the dynamics of cellular immune responses, which may be related to the severity of COVID-19. Third, at the time of convalescent plasma transfusion, the antibody level test (IgG, IgM et al) had not yet been routinely introduced at the hospital and no treatment guideline for using convalescent plasma was released. We did not measure the antibody concentrations in severe patients before and after convalescent plasma transfusion, so it is difficult to accurately evaluate the efficacy related to convalescent plasma.

In summary, all 18 patients with COVID-19 were discharged after treatment. Immunotherapy including convalescent plasma could be beneficial for severe patients. The strategies of early detection, early diagnosis, early isolation, and early treatment of COVID-19 in Qinghai are of importance to prevent the transmission and improve the cure rate.

## Data Availability

All data referred to in the manuscript are available.

## Acknowledgments

This study was funded by Science and Technology Department of Qinghai Province (number 2020-SF-158). We acknowledge all health-care workers involved in the diagnosis and treatment of patients in Qinghai. We thank all patients involved in the study. We also thank Xiangren A from Qinghai Provincial People’s Hospital for detection of coronavirus antibody.

